# Barriers and facilitators to alcohol support for South Asian communities: a qualitative framework analysis of UK service provider perspectives

**DOI:** 10.1101/2025.01.12.25320404

**Authors:** Stacey Jennings, Simon Dein, Graham R Foster

## Abstract

**Background:** Despite displaying pronounced alcohol-related physical and psychological harms, South Asian groups are critically underrepresented in alcohol treatment and research. Aggregate categorisation of diverse ethnic groups into ‘BAME’ collectives has perpetuated substantial knowledge gaps about the alcohol support needs of individual ethnic groups. Whilst there has been a recent growth in studies exploring the specific alcohol behaviours and support needs of South Asian groups, there is limited knowledge of professional experiences. This study aimed to address this gap by exploring service provider perspectives on barriers and facilitators to alcohol support for South Asian communities in the UK.

**Methods:** We carried out individual semi structured interviews with multidisciplinary staff spanning statutory and specialist services within East London in the UK. Interviews explored staff experiences and attitudes towards topics such as alcohol use triggers, maintenance factors, suitability of support options, and professional training needs. We adopted an intersectional lens during analysis to explore the influence and interplay of other relevant characteristics with ethnicity. We used qualitative framework analysis to identify relevant themes and map them onto a pre-existing risk environment framework across different levels (micro, meso, macro) of influence.

**Results:** 10 participants took part. 5 themes were developed that reflect barriers and facilitators to alcohol support across macro (‘service structure’), meso (‘cultural competence’, ‘gendered experience’ ‘religio-cultural norms’) and micro (‘lifestyle choice’) levels of influence. An intersectional lens indicated gendered, ethnic, and religious nuances in drinking and treatment seeking experiences. Whilst the overlapping nature of the micro-meso-macro levels was evident in the study findings, meso level factors were most established.

**Conclusions:** This study highlighted key areas of focus and unique barriers for diverse South Asian groups seeking support for alcohol misuse, with clear implications for culturally competent policy and practice in the UK context. Barriers such as short funding cycles, historical discrimination, ‘one size fits all’ approaches and training gaps on sensitive communication strategies pose challenges. Conversely, facilitators like targeted family education strategies and improving collaborative efforts between alcohol service types enhance support. Tailored specialist alcohol support for South Asian women is crucial.

## Background

Alcohol-related harms represent a long standing global public health challenge and heavy burden of avoidable harm(1, 2). Alcohol is a causal factor for a plethora of physical and mental health conditions, is associated with self-harm and suicide, and represents the biggest risk factor for death, ill-health, and disability among 15–49-year-olds in the UK(1, 3). The substantial societal costs are also well established, with alcohol estimated to cost the wider economy at least £27 billion annually across health, lost productivity and crime (2, 4).

Despite the UK’s alcohol strategy being underpinned by evidence-based NICE guidance and government initiatives for policy and practice(4–7), the broader picture is alarming. Alcohol-related mortality rates have reached an all-time high and treatment is characterised by low utilisation, high dropout, and substantial relapse rates(8–10). Criticism has been levelled at the governmental response, including the ‘particularly weak’(11) implementation of alcohol policy initiatives(12), as well as ‘blind spots’ in the evidence base regarding treatment barriers for ethnic minority subgroups (13–15).

The UK governmental Alcohol Strategy has not been updated since 2012(4). In the twelve years that have since passed, both alcohol-related harms and ethnic minority populations have significantly grown(9, 10, 16). Yet, ethnic minority groups remain underrepresented in standard treatment, overrepresented in alcohol harms and largely excluded in research(14, 17–19). Substantial knowledge gaps are perpetuated by aggregate categorisation of diverse ethnic groups into Black and Minority Ethnic (BAME) collectives, poor subgroup analysis and outcome reporting, and prevailing assumptions that underrepresentation indicates lower consumption (19). Reduced visibility of the alcohol support needs of individual ethnic groups makes the development of an appropriate response challenging. Likewise, whilst recently updated UK clinical guidelines for alcohol treatment highlight culturally sensitive approaches as a core element of practice(20–23), no centralised guidance exists and the complexities of operationalisation for specific ethnic groups has received little attention in alcohol treatment contexts (24). Encouragingly, recent government initiatives aim to address these limitations and improve ethnicity data standards by research with specific ethnic minority groups to find out ‘what works, why and for whom’(22, 25–27).

As the largest and fastest growing UK ethnic minority(16), the South Asian group represents a diverse collective of distinct subgroups (Bangladeshi, Pakistani, Indian and Sri Lankan). Each has unique religious and cultural traditions, further diversified by regional identities and generational differences. Alcohol occupies a complex and sensitive status across these groups whereby despite being religiously proscribed, cultural sanctioning and expression of behaviours is varied. The alcohol-harm paradox has also been observed whereby groups are overrepresented in certain alcohol-related disorders despite reporting low consumption (28–30). National treatment data (2022/23) also indicates a recent upward trend of people identifying as Muslim, Sikh and Hindu accessing treatment in England(8). Due to previously outlined inclusion, reporting and analysis limitations, reasons for this are unclear, but proposed explanations include delayed treatment seeking, community stigma, and cultural stereotyping by health professionals(17, 31–33). With a predominantly White British NHS workforce (74.3%)(34), much research has naturally focused on the influence of discrepant cultural factors and explanatory models (i.e., understandings of illness origins and trajectory) within the provider-user relationship on outcomes for South Asian groups(35–37). However, their applicability in an alcohol context is unclear, as are other salient social or structural factors, or how they interact.

East London represents one of the most ethnically diverse and rapidly growing regions in the UK, with multiple boroughs comprised of majority South Asian populations(38). East London also has some of the highest alcohol misuse, deprivation, crime rates and poorest health outcomes across England(39, 40). The considerable challenges local authorities face in developing tailored alcohol support for such areas necessitates examination of support needs at multiple levels. This study responds to academic and policy calls for an expansion to the lens through which we examine ethnicity and alcohol to consider relevant societal and structural factors(41, 42), as well as intersectional approaches that avoid spurious interpretations of ethnicity by considering multiple aspects of identity (e.g. age, gender, cultural background) (14). Despite a recent growth of intersectionality-informed research exploring the alcohol support needs of ‘BAME’ collectives (43, 44), to date there is limited knowledge on the specific needs of South Asian groups, or indeed on service provider insights for these groups.

This study aimed to address this critical gap by exploring provider perspectives on barriers and facilitators to alcohol treatment to help inform culturally tailored support for South Asian communities.

## Methods

### Design

Qualitative semi-structured interviews were conducted with 10 multidisciplinary permanent staff members from East London alcohol services and allied organisations, as well as specialist South Asian organisations accessible to East London residents.

### Setting

The study’s geographical parameter was delineated by the Bart’s Hepatitis C Virus Operational Delivery Network (Barts HCV ODN) which included the East London boroughs of Tower Hamlets, Newham, Waltham Forest, Hackney and Barking, and Havering & Redbridge. Study recruitment sites included National Health Service (NHS) alcohol inpatient and outpatient services, liaison teams, and community support services. Staff from relevant allied domestic violence and criminal justice services were also recruited, as well as established specialist South Asian organisations available to East London residents.

### Participants

A purposive sampling strategy was adopted to maximise variance of a broad range of occupational roles from diverse alcohol treatment pathways within the study perimeter. Inclusion criteria were: i) active or previous employment in a role routinely working with individuals with alcohol-related problems who identify as having South Asian (Bangladeshi, Pakistani, Indian) heritage and ii) willingness to be audio recorded. Sri Lankan groups were not included due to their significantly smaller proportion of the population in the East London study area of interest.

Staff at all sites were approached and asked if they wanted to take part. Snowballing methods were also used whereby participating staff circulated study information within their extensive professional networks to relevant colleagues who may be interested in taking part. All approached individuals took part in the study (n=10). Staff had an average age of 41 years old and five (50%) were men. Occupational roles were distributed across diverse pathways-details can be seen in Table 1. In line with updated government guidance and the Leeds Consensus Statement for ethical standards in ethnicity research(26, 45), staff were asked to self-ascribe their ethnic identity and that of the communities they typically work with. The average time in occupational role was 12 years (range 3-37 years).

**Table 1.** Study participant demographics (self-reported)

| **Characteristic** | **Participants** |
| --- | --- |
| **Sex** |  |
| Male | 5 |
| Female | 5 |
| **Time in role** |  |
| Mean (range) | 11 (4-37) |
| **Ethnicity** |  |
| Punjabi Sikh | 2 |
| Bangladeshi | 1 |
| British Asian | 1 |
| British Indian | 1 |
| Black African | 1 |
| White Polish | 1 |
| Afghan | 1 |
| White British | 2 |
| **Service type** |  |
| Primary care | 1 |
| Hospital | 1 |
| Community provider | 3 |
| Criminal justice sector | 2 |
| Domestic violence charity | 2 |
| Grassroots organisation | 1 |
| **Groups typically worked with** |  |
| All South Asian groups | 4 |
| Punjabi Sikh | 2 |
| Bangladeshi | 3 |
| Muslim and Hindu | 1 |

### Procedure

Ahead of interview, participants were provided with an information sheet detailing the aims of the study. The lead author arranged a pre-meeting to discuss the research in full, answer questions, and confirm inclusion/exclusion criteria. Interviews were then arranged at a time and location convenient to the participant and informed e-consent was taken. Flexibility in interview method (i.e., in person or remote) was offered to balance participant preference and the changing status of COVID-19 related restrictions throughout the study. All participants were interviewed using video conferencing software (Microsoft Teams), audio recorded with a password protected Dictaphone and transcribed verbatim. Participants were offered a £10 Love2shop voucher for their time. Recruitment continued until theoretical data saturation was reached(46–48), i.e., when a rich analysis had been obtained and subsequent interviews yielded no new information.

### Data collection

Data was collected from January 2022 to January 2023 via individual interviews which lasted between 31 minutes to 65 minutes (mean=47 minutes). A draft topic guide was developed based on key themes identified in a previous literature review. Two PPI groups reviewed the topic guide, which was then pilot tested and minor edits made (see Appendix 1). During interviews the topic guide was used flexibly with attention given to any participant-raised issue. Audio recordings were immediately uploaded to secure password-protected servers post interview and deleted once successfully transcribed. To preserve anonymity, participant names were replaced with appropriate pseudonyms derived from the BabyNamesDirect database^242^. Any additional identifiable information provided in analysis excerpts were also anonymised. Leeds Consensus Statement guidance was followed throughout to promote ethical practice and avoid perpetuating ethnic stereotypes(45).

### Analysis

Interview data was transcribed verbatim and imported into NVivo v12 software for line-by-line coding. As this study aimed to identify barriers and facilitators across multiple levels, Rhodes risk environment(49) was selected as a suitable theoretical framework as it was devised specifically for substance misuse contexts to identify harms across individual (micro) factors, as well as wider group (meso) and structural (macro) mechanisms and their interactions. Data was analysed using framework analysis, a highly appropriate analytic strategy as it permits explicit mapping of themes against pre-specified categories across the risk environment(50, 51). Framework analysis also permitted explicit exploration of intersectional characteristics on drinking and help seeking behaviours.

The five steps of framework analysis were followed: i) familiarization with data, ii) identifying a thematic framework, iii) indexing of themes, iv) charting of themes within the framework, and v) mapping and interpretation. As per previous studies, reflexive thematic analysis(52–54) was incorporated at indexing and charting stages to inductively generate themes and subthemes from participant accounts which were then mapped onto the pre-existing deductive (micro, meso and macro) categories. Noble’s credibility checklist was implemented to enhance the credibility of findings(55). This study has also been reported in accordance with the Standards for Reporting Qualitative Research framework (SRQR)(56) to improve transparency and reporting standards.

## Results

A total of 5 themes and 10 subthemes were identified across macro, meso and micro levels. Each major theme and its constituent subthemes are discussed in depth below, and conceptually depicted in Figure 1.

**Fig 1.**
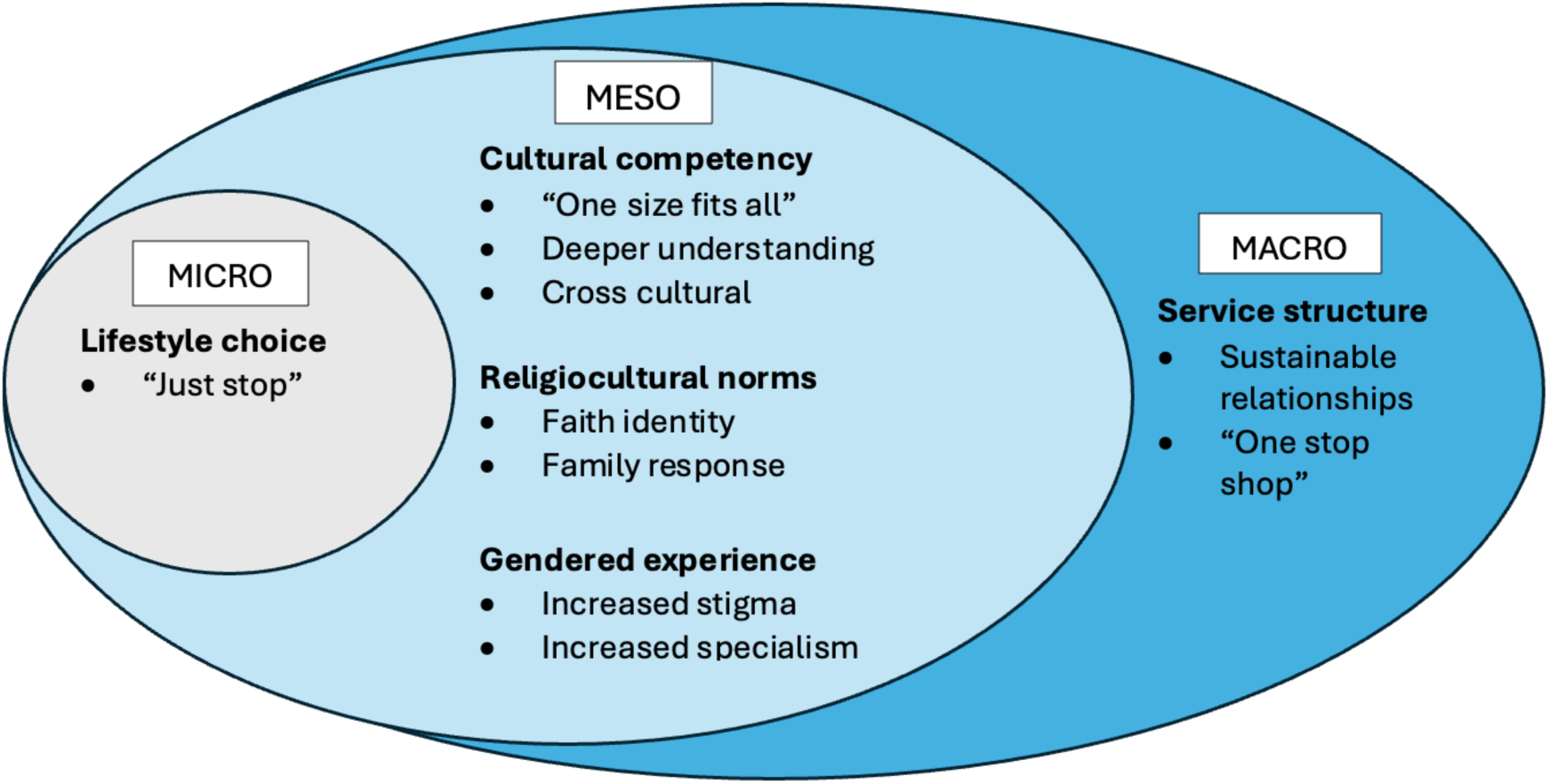
Conceptual framework of provider barriers and facilitators

### 1. Service structure

#### a) Sustainable relationships

Described as a “never-ending problem”, decommissioning and funding restrictions were seen to have a detrimental impact on both the accessibility and quality of targeted alcohol support. Staff believed short funding cycles practices undermined the vital development of long-term collaborations between services and key community figures (including faith leaders).

> *Agata: “Because if there was enough funding and time then we could reach out to those communities. So you can actually go to Imams and speak to them, or the Imans could give a sermon or they can engage with the family together if someone is struggling to access treatment.*” *(Polish, criminal justice pathway)*

Many staff highlighted decommissioning of the few reputable South Asian-specific alcohol services as a concern, with grassroots organisations increasingly attempting to fill this gap in cultural peer support. For one participant this was reflected in the “incredible popularity” of his organisation’s Punjabi male support groups, reaching folks “dealing with things that my non-Punjabi friends wouldn’t understand.” Several staff members also described how a lack of funding curtailed the ability of services to resolve early implementation ‘teething issues’,

> *Rehan: “… Sure it brought a lot of challenges, but that might have been a bit hard to overcome those challenges with such a short funding cycle.” (Punjabi, grassroots organisation)*

Two providers suggested long-term borough wide initiatives to combat these problems, with one detailing how her service reflected a “complete successful worked example” by developing a forum of South Asian staff to build relationships with diverse local religious governing bodies.

> *Ashi: “…go right to the top and build with the actual Sikh organisations that govern the area, so that even if a committee does change, on a national level, they are aware that we are working with them. Then Sikhs (like myself) could talk to Gurdwara and Hindu members of staff to talk to Hindu organisations to build that rapport on a national level and filter it down…” (Punjabi Sikh, community provider)*

Several participants described the need to improve working relationships and address existing tension between service types. One provider described how an ‘us vs them’ divide was frequently perpetuated in public discourse between mainstream and grassroots services, which felt like “shooting the entire community in the foot”. Another described the challenges of sharing treatment effectiveness data between hospital and community providers as “nigh on impossible”.

> *Ashi: “There some awesome individuals out there right now who are ex-alcoholics highlighting alcohol within our community, which is absolutely fantastic. I mean, any awareness is good, but then because they’re using it to say mainstream services don’t help you and they’re not there for you” (Punjabi Sikh, community provider)*

> *Thea: “We need to know if it’s working, they need to be analysing the options that they’re offering patients because if it’s not working, we need to rethink…” (White British, medical doctor)*

Participants from grassroots and community-driven organisations felt that integral to improving relationships was recognition of their contribution, an avoidance of tokenistic and extractive approaches, and the development of reciprocal partnership working to “share resources, knowledge and expertise”.

> *Rehan: “If we’re not invested in the community organisations, the only thing that we want from them is their work and their reach- we don’t actually want them to last.” (Punjabi, grassroots organisation)*

#### b) “One stop shop”

All participants discussed recent structural shifts within local authorities to a community “one stop shop” integrated alcohol and drugs model. Whilst one participant advocated for this model as having the mainstream “firepower” to support specialist approaches, other participants were largely critical of the multiple barriers it enacted by limiting choice for service users and reducing ability to meet specific local need.

> *Diyan: “The other part is flexibility- so, what you find is that a lot of the tenders for drug and alcohol services go for things that are just a blueprint of what they set up for others. They have like a template or business model and a lot don’t really do their research into the demographics of a specific area, so then don’t propose specific niche interventions for a particular community”. (Bangladeshi, community provider)*

The integrated model contributed to the perception of a generic ‘West-centric model’ that predominantly served White British service users. Clients were perceived as being “funnelled” through a rigid ‘White’ way of treatment that didn’t align with user preferences or experiences; particularly group therapies which were deemed significantly less acceptable to South Asian communities than their White British counterparts.

> *Diyan: “I think the way the treatment is designed, it’s very perhaps, Western orientated, very focused around sharing and being open about your feelings and looking at recovery goals, etc. And that’s not the case with South Asian communities, you know, talking about your personal life, it’s not something that you do, especially in a group of strangers, the guilt and shame, all those things are attached.” (Bangladeshi, community provider)*

Furthermore, the integration of drugs and alcohol service components under this umbrella service was perceived to compound stigma for South Asian groups, with one participant expressing a strong preference for alcohol-specific services.

> *Thea: “I think because they lump drug and alcohol together there’s a whole stigma of “well, we’re not druggies”. It hasn’t worked, putting those two services together as substance misuse services. You need to have a variety of help for patients”. (White British, medical doctor)*.

One participant felt historical relationships were a fundamental yet often overlooked barrier in explaining disengagement between South Asian communities and alcohol services.

Generalisation of statutory services contributed to a perception of proximity to the state and being unsafe. Conversely, grassroots organisations were perceived as being “more representative of their communities, with more transparency and accountability”.

> *Rehan: “…those types of experiences are rarely considered within the space of mental health, alcohol… As soon as you have a service that’s perceived as proximate to the state, I think it has an impact on trust.” (Punjabi, grassroots organisation)*

Nonetheless, all staff recognised the importance of specialist approaches in to successfully engage and address alcohol problems in South Asian communities.

## Meso level

### 2. Cultural competence

This theme captured key areas of cultural sensitivity providers identified as requiring improvement, as well as nuanced (and occasionally conflicting) perspectives of how these should look in service delivery.

#### a) ‘One size fits all’

Participants described a treatment gap whereby despite South Asian communities being aware of available services, they were still unlikely to engage. One participant described this as reflecting the distinction between equity and equality, with alcohol and mental health services “not meeting folks where they are”.

> *Rehan: “Just because everyone has equal access to services doesn’t mean that they are starting from the same point of process to actually understand or even get involved with services…” (Punjabi, grassroots organisation)*

> *Daniel: “They get referred to services but then they don’t attend or participate in groups, so they become hard to reach and to help.” (Black African, hospital provider)*.

This was attributed to a ‘one size fits all’ approach unable to adapt to the diverse needs of South Asian communities. All participants specifically focused on staff demographics and training. While some believed ethnic matching of staff and service user characteristics was essential for successful engagement, others highlighted stark variation in service user preference. Many participants shared experiences of clients explicitly requesting to “speak to someone who isn’t from their background*”* due to fear of gossip in the community, which confidentiality reassurances did not override.

> *Sally: “…it’s quite difficult to make it clear that you’re a professional, that you’re talking as someone who’s supporting them rather than talking as someone, who’s a) going to gossip or b) disapprove of them…”. (White British, primary care)*

Providers also described significant challenges surrounding balancing language and confidentiality needs. Whilst translation services were recognised as valuable, several participants described the presence of an interpreter as a “massive barrier” by altering the dynamic of therapeutic sessions.

> *Diyan: “They haven’t built a rapport and all of a sudden you’re expecting them to translate real kind of traumatic experiences of their life.” (Bangladeshi, community provider)*

Participants agreed that whilst “not 100% perfect” to address confidentiality concerns, a pragmatic solution would be to offer flexibility in choice to the service user (interpreter *or* staff member speaking same language). However, participants robustly recognised the challenges imposed by funding and resource limitations in accommodating service user preference, particularly in restricting team diversity and capacity.

> *Agata: “I’ve got a really tiny team so women will just see me. That’s it. I’m the only choice.” (Polish, criminal justice pathway)*.

#### b) Deeper understanding

Several participants felt that shared backgrounds didn’t necessarily confer cultural competence. Instead, the key component was a deep understanding of wider South Asian historical and sociocultural contexts, whilst simultaneously not assuming how this relates to the individual.

> *Rehan: “Cultural competence in my mind doesn’t just mean that folks from particular backgrounds are represented in services, but it’s almost like the understanding that someone within a service doesn’t have to explain their whole upbringing. It’s more about people who are open to listening and not making assumptions or projecting their assumptions onto services users” (Punjabi, grassroots organisation)*

This extended to the “essential need for an intersectional approach” that also considers subsections within South Asian communities (such as LGBTQIA) that may “require more targeted or more specialist support”.

> *Rehan: “It’s recognising that it might be some folks who, for example, may identify as a Punjabi man but then also identify as queer. And then being responsive to how we can develop support for those who we may be unintentionally missing out”. (Punjabi, grassroots organisation)*

Training was seen as an essential component to facilitate deeper understanding. Several participants described how approaches must expand beyond narrow focus on stigma and harmful cultural practices to include the relevant historical and structural “nuances and layers” for contextualised understanding.

> *Khadija: “I think a lot of people feel like once they’ve understood honour-based violence and forced marriage and FGM [female genital mutilation] that’s it-they got some training on that so they’re culturally competent, now they know it all!…immigration, underemployment, class structures, systemic oppression- to be having a deep understanding of that means that you’re actually culturally competent” (Afghan, domestic violence charity)*

> *Rehan: “I’m getting so tired of seeing stuff on stigma and mental health in Punjabi communities, like, we’ve moved forward from that.” (Punjabi, grassroots organisation)*

One participant also discussed the need to frame cultural competence as a continual learning process to align with constant evolution of cultural and healthcare behaviours.

> *Khadija: “Having specific policies, constant reviews of how staff members are doing and sort of, taking it as a learning journey rather than an end goal I think to me is being culturally competent. You must be committed on so many different layers.” (Afghan, domestic violence charity)*

However, many participants described stark inadequacies in training provision as “just a tokenistic approach of delivering training on EDI*”*, or an absence of formal training altogether. Two participants highlighted guidance on sensitive communication strategies during alcohol screening and *“*getting staff feeling comfortable talking about these issues*”* as a specific training gap.

> *Sally: “For example, we run GP alcohol certificate training, but there’s no part of that that’s actually practising what you say for example. We are encouraging staff to do AUDIT-C, but there’s no guidance…?”. (White British, primary care)*

#### c) Cross cultural factors

Participants stressed that certain barriers and facilitators were shared irrespective of service user ethnic background. The “chaotic” environment and unpredictable nature of service waiting rooms was a commonly cited cross-cultural barrier.

> *Prit: “So someone who’s only using alcohol, like a young person just using alcohol or cannabis maybe, would they feel comfortable with older heroin users and crack users and coke users?” (British Asian, criminal justice pathway)*

Resistance to engaging with support whilst overwhelmed with alcohol-related problems was seen as a widespread issue. Staff also noted that moral tension and social repercussions regarding drinking exist within families in most subsections of UK society. Likewise, the widespread accessibility and advertising of alcohol was thought to undermine preventative efforts and reinforce acceptability for everyone.

> *Diyan: “I think this is just society as a whole, I don’t think that the dangers of alcohol are highlighted enough because it’s still an open and accessible thing advertised and widely accepted.” (Bangladeshi, community provider)*

Lastly, participants recognised that discretion of services to remove stigma was a shared facilitator, typically via embedding into neutral health spaces and benign naming that avoids explicit “labelling as an alcohol service”.

> *Ashi: “That really, really helps when you have links in your borough to GP surgeries to the Gurdwara to the community spaces, so that when people from South Asian communities, be it Muslims Sikh Hindu to the outside eye, they’re just going to the doctor, going to the Mosque…”. (Punjabi Sikh, community provider)*

### 3. Religio-cultural norms

#### a) Faith identity

All participants described the differing acceptability of alcohol use (and subsequent family/wider community responses) across subgroups, shaped by intersecting religio-cultural traditions and gender. Providers described how alcohol proscription was most salient within Islam and referenced a ‘hierarchy of acceptability’ which placed Muslim as the least accepting of alcohol (particularly in relation to women-discussed in the next theme) and Sikh as the most accepting.

> *Diyan: “when I look at the South Asian community, I divide it into the different religions. So you have Islam, you have Hinduism, and then you have Sikhism, and then that’s when I get the context of how the alcohol is consumed. What I find is that alcohol is quite a social thing in the Sikh community and it’s predominantly male orientated.” (Bangladeshi, community providers)*

Alcohol was seen as even more taboo or ‘haram’ than illegal drugs in Muslim culture, with one participant likening it to a White British counterpart “pulling out some foil to take heroin in front of everybody else”. Despite this, staff observed wide variation in alcohol behaviours. Whilst several perceived this as natural expression of differing interpretations of religious texts, others questioned their legitimacy and strongly believed any association with alcohol diminished religious status.

> *Sally: “it is quite complicated. And it’s all quite mixed and there’s a huge diversity in what people think is acceptable and what people think the norm is”. (White British, primary care)*

> *Diyan: “So even if you sell alcohol, then it’s the same sin as drinking it. So now there’s another shift happening where if you look at all the Indian restaurants, they are predominantly owned by Muslim Bangladeshi owners and that concept hasn’t obviously kicked in with them yet.” (Bangladeshi, community provider)*

Reframing the association between faith identity and drinking was seen as fundamental to allay fears about jeopardizing one’s religious status, reduce stigma and promote help seeking. Several staff outlined successful collaborations with places of worship to promote alcohol support within communities, with compassion-led approaches seen as a particularly effective strategy.

> *Rehan: “I think what I’d like to focus on is also potential guilt in relation to folk’s faith identities, as well as their relationship with alcohol. There is such an important part in this around reframing how folks relate to faith practise, and rather than it being like, “oh, if you drink, that’s it, you’re a bad Sikh (whatever that means!”), but it being a bit more compassionate.” (Punjabi, grassroots organisation)*

> *Prit: “I think the Imams doing a lot of promotional work and then trying to educate people that we shouldn’t be judgemental, and we should try to support whoever is in the community.” (British Asian, criminal justice system)*

Providers also described how the embedded nature of faith beliefs within South Asian communities distinguished them from their White British counterparts. As such, the religious underpinnings of common treatment modes (like Alcoholics Anonymous) could “clash” with existing beliefs and be particularly offputting.

> *Agata: “Generally speaking a lot of South Asians definitely struggle with the concept of 12 step…it is such a clashing because it’s this other book and religion and they’re supposed to follow their own religion.” (Polish, criminal justice pathway)*

Another participant described how this embedment often manifested during alcohol screening and outlined neutral framing methods to combat this ‘deference to religion’.

> *Sally: “…what you often get as a reply if you just ask the family or individuals or whatever is “Oh no, I’m a Muslim” so I almost say, “I don’t want you to feel offended but I’m asking you about this because I have to ask questions”. (White British, primary care)*

#### b) Family response

Family responses were primarily driven by stigma from bringing shame via contravening religious and sociocultural norms. Encouragement to conceal drinking problems within the family contributed to delayed treatment seeking and increased severity of clinical presentations. Mirroring the hierarchy of acceptability discussed in the previous theme, participants described a hierarchy of stigma with Muslim groups occupying most stigmatised position. Gossip was recognised as the central perpetrator across all subgroups, described as the South Asian equivalent to “CCTV camera surveillance”. One participant described how even working in the alcohol sector carried significant stigma, illustrated by how rapidly gossip spread within her community:

> *Ashi: “I remember when I started working in substance misuse services and my first day of work, I got home and my dad goes “by 9:30 I got a phone call saying that you’ve got a drug problem”… because traditionally, we should be doctors or employees or pharmacists or something like that, so it’s like why are you working in the drug and alcohol sector? It’s a stigma from both ends…” (Punjabi Sikh, community provider)*

> *Agata: “I’m thinking about Bangladeshi, and they’re usually from Muslim families, and alcohol or drinking is not looked at as good at all… So before someone actually gets to us to get support, things might get really bad before they actually access support and really their drinking is quite risky”. (Polish, criminal justice system)*

Staff had varied perspectives on the benefits of family involvement for an individual’s treatment outcomes. Families were often perceived to only approve of a limited number of treatments (typically medically-based detox) and “doubt or don’t understand the point of therapies or recovery workers” with some encouraging exclusive reliance on religion or community alone. One participant also described how family consultation in primary care could inhibit discussions around alcohol.

> *Sally: “So behind the curtain you may want to say look I need to examine you now and then you have to go (motions drinking to patient) but it depends on the person, you see.” (White British, primary care)*

> *Ashi: “So to tap into the Muslim community has been harder for us, because it’s like no. It’s not for drug agencies. No, Allah will help you and the community will help you-you don’t need to take it out the community”. (Punjabi Sikh, community provider)*

However, staff perceived the often-larger support network of South Asian individuals as a unique resource filled with potential. Unfortunately, the effects of stigma and concealment contributed to a cycle whereby immediate social circles were deemed ill-informed and ill equipped to help. Staff identified specific gaps in family alcohol literacy including insufficient understanding of the harms beyond liver damage, including the association with wider community issues such as domestic violence and crime.

> *Diyan: “So I think the education around the harms of alcohol, I don’t think it’s been promoted enough within those communities. And I think that’s solely because of the whole kind of stigma. And so, these people are less likely to seek help unless the doctor comes and says you need to go and check your liver out etc”. (Bangladeshi, community provider)*

Conversely, others felt the primary knowledge gap to be addressed was appropriate communication with someone struggling with alcohol. Parallels were drawn with mental health conversations in the community.

> *Rehan: “This is related to alcohol, but also other areas, especially when we talk about mental health, one of the biggest questions that we get is-how do you raise this topic? Because unless we have the communication or the language to be able to raise it, we’re unable to kind of name these difficulties and potentially take steps forward…” (Punjabi, grassroots organisation)*

Staff also highlighted the need to expand alcohol campaign audiences beyond those already dealing with problems to include families and lay people in the community. The prominence and perceived respectability of the educator was also deemed to be crucial, with one participant deeming “you can’t get higher than doctors in our community”. One participant described the respective benefits of a medical approach compared to a more emotive approach rooted in lived experience.

> *Ashi: “Recently, they did a session in the Gurdwara and an ex-alcoholic came in and spoke about it. It really held the room…So the medical side can tell you exactly what it does but when it’s a person sitting in front of you and saying to you, this is my journey, this is how it screwed up my life, but I beat it and I’m here". (Punjabi Sikh, community provider)*

Collaboration with faith leaders was commonly highlighted as a highly effective method of family and lay community education.

> *Diyan: “In Tower Hamlets, they had a drug and alcohol worker based in the Mosques. Their main role was to ensure that, firstly, they trained all the Imams around alcohol awareness and talk about the dependency side of things so when somebody comes in, we don’t tell them just to stop straight away. It was important to tell them about the stages of somebody’s recovery and what they might do, and where they can be referred etc”. (Bangladeshi, community provider)*

### 4. A gendered experience

#### a) Increased stigma

All participants considered South Asian women as a distinct group facing unique barriers and facilitators to alcohol support. Unlike the intersectional variations observed for males, women’s drinking was strictly unacceptable and taboo across all South Asian subgroups. However, due to religious proscription Muslim women were seen to be particularly stigmatised.

> *Ashi: “Culturally it’s okay for some Punjabi males to drink but culturally, if you’re Punjabi woman, who drinks, no. The shame attached to that!” (Punjabi Sikh, community provider)*

> *Khadija: “Whereas I would say that Indian women are probably more likely to talk about it and more likely to seek support as opposed to Pakistani or Bangladeshi or women from other Muslim countries in the region”. (Afghan, domestic violence charity)*

Stigma was seen as the fundamental driver for ineffective community handling of women’s drinking. This gendered sense of shame was perceived as being deeply engrained at an early age and associated with family respect and traditional caregiver roles, fundamental principles which guided behaviour.

> *Ashi: “In our culture we’ve got a saying: “izzat jaan to pyaari”. It’s roughly translated as your respect is worth more than your life. Death before dishonour basically. In our culture women are seen as the respect of a household, so if you as the woman lose respect, the whole household loses respect… we have we have that engrained into us from the day we are born.” (Punjabi Sikh, community provider)*

Providers described the typical clinical presentations of South Asian women as including complex features such as multiple substance use, adverse childhood experiences, social isolation, and interpersonal violence. Relationships with partners were frequently implicated in drinking behaviours, with one staff member hypothesising that isolation was a possible cause or result of heavy alcohol use. Whilst avoiding external help and concealing drinking problems was perceived as the social expectation, unlike men, in some extreme cases women even concealed from their immediate families. Commonly observed family responses to female drinking were perceived as inadequate or inappropriate, such as providing the woman with additional family or employment responsibilities. Another participant described the “double stigma” women faced when seeking help for alcohol use in the context of domestic violence.

> *Khadija: “… if you’re using alcohol or drugs as a coping mechanism as a result of abuse, then disclosing that and seeking help for that is just an extra layer of burden. So the solution is always not to seek help because it’s a better alternative than having to go seek help”. (Afghan, domestic violence charity)*

#### b) Increased specialism

Providers robustly expressed that the entrenched stigma and hidden nature of women’s drinking requires “a lot of specialism” and that some initial resistance to engagement should be expected.

> Ashi: *“I think it’s gonna be slower to engage South Asian women. Especially Muslim women. It’s definitely a big no-no there and it’s gonna take a lot longer to get that one kind of at the forefront and but it is a real issue”. (Punjabi Sikh, community provider)*

Staff identified critical features of cultural competence to improve engagement, including the need to acknowledge diversity and avoid treating South Asian women as a monolithic group. Strikingly, several staff highlighted how alcohol and allied services could endanger women through the inadvertent normalisation of ‘cultural’ harmful behaviours and inappropriate language. Poor experiences within alcohol services often had detrimental knock-on effects for engagement with allied support services such as domestic violence agencies.

> *Khadija: “A lot of times the term cultural sensitivity is used to be sympathetic towards perpetrators and culture is used as an excuse for domestic abuse. You have to be very careful what terminology you’re using… Even if they were to go and seek support, that support in itself may alienate them even further… once they are referred to drug and alcohol support, they slowly stop engaging with domestic violence services as well or stop engaging with all services completely.” (Afghan, domestic violence charity)*

Unlike their male counterparts, direct involvement of religious leaders in alcohol support was also seen as being potentially harmful for women.

> *Khadija: “Especially because of honour based violence, you have to be really careful with how you frame it. I think it could be very very dangerous and could put a lot of women at greater risk of harm than could do good”. (Afghan, domestic violence charity)*

Many participants also expressed frustrations at the lack of availability and focus on dedicated services for women, which were typically treated as a “secondary consideration”. One staff member expressed difficulty locating any specialist support for a female client.

> *Daniel: “[It] would be something very useful to have given the unique problems that they all seem to share… I remember going on to this website to see if I could find something more appropriate, and I couldn’t. It’s just about maybe something more specific could make the difference”. (Black African, hospital provider)*.

Some providers even deemed mainstream services unsafe in their current form, and advocated for the provision of designated women’s only safe spaces until standard services were culturally equipped to provide the level of specialism required.

> *Khadija: “So until we get there where mainstream services can be 100% safe for people who face multiple barriers then I definitely advocate for women only spaces or, like POC [People of Colour] only spaces because of that very reason…I think it would help a great deal to provide those spaces for women who are experiencing substance dependency”. (Afghan, domestic violence charity)*

## Micro level

### 5. Lifestyle choice

#### a) ‘Just stop’

Participants perceived that wider South Asian communities often framed alcohol-related problems as a conscious personal decision “entirely down to their choice around lifestyle*”*. This directed attributions of blame and responsibility solely to the individual, enhancing stigma and contributing to a belief that motivation to change should be sufficient to ‘just stop’. This could deter people from seeking external support outside the family unit or religious circle, reinforcing the perception that continuing to drink was an active choice. Often the dangers of suddenly stopping drinking were not well understood.

Staff believed increased education around the nature of addiction would help shift these perceptions by providing a more “compassionate and contextualised” understanding and better equipping family and community members to support those affected.

> *Rehan: “Oftentimes the response is a lack of understanding… And I feel what we’ve seen in our work as well is that when people don’t know how to confront a particular situation, oftentimes we kind of default to a response which is usually quite stigmatizing and blaming the person for the difficulties they’re going through.” (Punjabi, grassroots organisation)*

## Discussion

This was the first study to examine the perspectives of UK service providers on barriers and facilitators to alcohol support for South Asian groups. Over half (n=5) of participants were from South Asian backgrounds and naturally drew heavily on personal experiences as well as professional in their accounts.

The overlapping nature of the micro–meso–macro divide resonates in the study findings; however micro level factors were less established. A single theme ‘lifestyle choice’ described how harmful perceptions of individuals being solely culpable for alcohol problems reduced external help seeking in South Asian groups. Such conceptualisation of addiction as a conscious ‘failure of will’(57) has been a long-contested subject of national debate. Whilst advances to formally recognise addiction as a classified mental health disorder have helped shift public perception(57, 58), persisting attitudes that those with alcohol dependence are more responsible for their condition may be magnified in South Asian communities where alcohol is highly stigmatised(59). Targeted family educational campaigns surrounding the nature of addiction may facilitate change in such beliefs.

Recently published UK clinical guidelines for alcohol treatment promote an ‘inclusive service ethos’ that requires services to be culturally competent and tailored to meet local need(23), promoting team diversity and staff training as key facilitators (60). However, ‘meso’ level findings in this study highlight nuance within these specific areas that must be considered to develop truly inclusive services. Firstly, providers indicated that models in their current ‘one size fits all’ state were unable to adapt to local need, rejecting persistent beliefs that underrepresentation reflected poor awareness of available alcohol services. Likewise, whilst staff diversity is undoubtedly beneficial, ethnic matching of staff and service user can be a particularly contested topic for South Asian engagement(61–67) and should not be assumed as a default approach. Study findings also mirror cautionary recommendations to balance a sensitive understanding of South Asian traditional and historical contexts without assuming individual preferences (23). The challenge is how to successfully reconcile these concepts in clinical practice (68, 69).

Whilst staff training is fundamental to cultivating such understanding, this study highlighted significant gaps in provision and content. Many providers highlighted a complete lack of formal training with overreliance on existing staff knowledge, risking further stereotyping marginalised groups(70, 71). Participants also suggested several components to enhance cultural competency training, including sensitive communication strategies, incorporation of societal and structural contexts, adopting intersectional approaches that consider the needs of community subsections (e.g. LGBTQIA+), and framing training as an ongoing process rather than an ‘end point’ Likewise, whilst the impact of interpreter use on confidentiality and therapeutic dynamic shifts has been well documented(72–74), it is unclear if previous suggestions such as hiring minority staff outside local communities can sufficiently allay such profound fears in the alcohol context(63). Feasible solutions to the conundrum of accommodating both privacy and language needs are clearly required.

Corroborating previous research, nuanced intersections between religiocultural traditions and gender roles shaped drinking behaviours, responses, and support needs. Providers recognised a broad alcohol ‘hierarchy of acceptability’ based on religious proscription, where stigma was more salient among Muslims and Asian women generally(17, 73–75). Consideration of the sensitive and culturally embedded nature of faith beliefs in South Asian communities is paramount to avoid discord, moving away from the clashing religious underpinnings of common treatment modalities in favour of compassion-led approaches that address religious guilt. Staff recognised large family support networks as a potentially valuable resource, undermined by a vicious cycle of concealment leading to poor alcohol dependence literacy.

Echoed by previous studies, family presence could also impede disclosure in primary care settings and reject non-medical treatment options(76). Sensitive communication strategies surrounding alcohol in both informal (familial conversations) and professional contexts (alcohol screening) were identified as a core area of need. As family involvement is a core principle of care, providers should be sensitive to the nuances of existing stigma when developing tailored support for individuals and their families.

Whilst previous East London based research positioned faith leaders as an effective agent to reduce stigma and normalise help seeking(68), this study identified gendered distinctions for South Asian women reflected across unique triggers, help seeking and support needs. Unlike their male counterparts, the involvement of faith institutions was seen as potentially harmful. Findings of complex clinical presentations defined by adverse childhood experiences, isolation, domestic violence, and multiple substances corroborate those in Galvani et. al’s (2023) recent report (73), yet a lack of relevant treatment data makes it difficult to compare this profile with women of other ethnic backgrounds (77). However, closer partnership working between domestic violence and alcohol services may be valuable to facilitate early identification and concurrent treatment across services (78). Caution around misinformed cultural sensitivity approaches that inadvertently place women at further risk of harm was highlighted as a key priority, specifically the use of inappropriate language or ‘normalising’ abusive behaviours, which could trigger widespread service disengagement(77, 79).

Additionally, engagement strategies deemed appropriate for men cannot be assumed to be effective, or even safe, for women (78). This underlines the fundamental importance of carefully defining and operationalising cultural competency with an intersectional lens within alcohol contexts. Further research is urgently needed to inform specialist approaches and adequately tailor alcohol services to meet the unique needs of South Asian women.

At the macro level, service restructuring and funding practices curtailed effectiveness and engagement with South Asian communities. Echoing previous research(80, 81), short funding cycles inhibited the development of sustainable relationships with communities and opportunities to resolve early implementation challenges. Likewise, structural shifts towards an integrated community model enhanced perceptions of being a state-proximate service and imposing unacceptable ‘White’ ways of treatment (such as group-based therapies).

Conversely, grassroots, and ethnic minority organisations were perceived as transparent and trustworthy. As recognised as a national priority by the NHS Race Observatory, improving trust between mainstream services and ethnic minority communities, as well as across different service types, should be a key area of focus to establish co-ordinated, reciprocal collaborations and improve outcomes(60, 82). In the context of recent additional investment for alcohol treatment, local authorities must ensure they involve local communities and organisations to co-produce services tailored to meet diverse local need.

### Strengths and limitations

To the authors knowledge, this is the first study to explore staff perspectives on barriers and facilitators to alcohol treatment specifically for South Asian communities. Inclusion of a considerable range of professional voices ensured findings reflected the wider alcohol workforce. This study also responds to calls to adopt broader intersectionality-informed approaches that expand beyond individual levels of influence, as well as distinguish South Asian-specific factors to avoid contributing to an unsophisticated understanding of ethnic groups that perpetuates stereotyping (14, 70). Application of the risk environment framework enabled barriers and facilitators to be clearly mapped across micro, meso and micro levels, and their complex interplay explored. This is also one of few UK studies that has addressed the alcohol support needs of South Asian women- a critically underexplored issue. Likewise, findings across micro, meso and macro levels directly inform key priorities and principles of care outlined in the recently published UK clinical guidelines for alcohol treatment.

This study has some limitations. Firstly, participants were recruited from East London boroughs within a pre-defined operational network. Findings cannot be assumed to uniformly apply for all South Asian communities due to differing social, economic, migration and cultural contexts, as well as large variation in local authority funding and provision across locations. However, qualitative studies can still uncover rich insights into pertinent health and social issues that make a significant contribution to policy and practice(83). As such, study findings should be primarily interpreted in the context of East London, with cautious broader application made to the rest of the UK where appropriate. Likewise, as alcohol cultures and ethnicity are dynamic concepts and not ‘static’ units of analysis, future research exploring generational experiences could further illuminate the alcohol support needs of South Asian communities across the UK.

## Data Availability

The dataset generated during this study are not publicly available to maintain confidentiality agreements and avoid potential identification of involved organisations and individuals. Contact the corresponding author for further information.

## Declarations

### Ethics approval and consent to participate

This study received approval from Queen Mary University of London internal committee and Camden and King’s Cross Research Ethics Committee (21/LO/0337). Informed consent was taken from all study participants.

### Consent for publication

Not applicable.

### Author contributions

SJ led on conceptualisation of study design, data collection, analysis, and interpretation, and manuscript writing. SD and GRF jointly supervised the study and contributed to data analysis, interpretation, and manuscript drafts. All authors provided feedback on the draft manuscript and approved the final manuscript.

## Acknowledgements

We thank our participants for taking part in this research, as well as our stakeholder groups for their input in the design and interpretation of the study. We also thank the alcohol service networks within East London for supporting the study and Barts Health Charity for funding the study.

